# Maternal prenatal stress during the first trimester and infant birthweight in Soweto, South Africa

**DOI:** 10.1101/2021.01.11.21249590

**Authors:** Andrew Wooyoung Kim, Rihlat Said Mohamed, Christopher W. Kuzawa, Shane A. Norris

## Abstract

**Background:** Approximately 14.2% of newborns are estimated to be born low birth weight (LBW) in South Africa. Past work has implicated maternal prenatal stress as a potent predictor of poor birth outcomes, including preterm birth, intrauterine growth restriction, and LBW. However, less is presently known about the impacts of prenatal stress during early gestation in low- and middle-income contexts, where public health burdens due to LBW are much higher.

**Objective:** We assess the effects of psychosocial stress during the first trimester on birthweight in a large sample of women (n = 657) in Soweto, South Africa, a peri-urban township located in the Greater Johannesburg area.

**Methods:** Data come from the Soweto First 1000 Days Cohort, a study of maternal and fetal predictors of infant birth outcomes. Multiple regression models were used to examine the impact of prenatal stress on infant birthweight.

**Results:** The prevalence of LBW was 16.6%. Adjusting for maternal age, gestational age, fetal gender, body mass index, and parity, maternal prenatal stress during the first trimester was a borderline predictor of lower birthweight in this sample (β = 12.7, *p* = 0.071, 95% CI [-26.4, 1.10]). Women who reported greater levels of stress appeared to have non-significantly longer gestations, and the negative impact of maternal stress on birthweight only appeared after adjusting for this pattern.

**Conclusions:** These results suggest that fetal growth restriction associated with first trimester maternal stress may contribute to lower birthweights in this sample. Our findings report modest relationships between maternal stress specific to early gestation as likely important to birth outcomes in this urban South African sample.

## INTRODUCTION

Being born low birthweight (LBW, BW<2500g at birth) increases one’s risk for a wide range of poor physical and mental health outcomes that can persist across the lifecourse, and can even have effects that impact health in the next generation (Barker 1998; Kuzawa & Sweet 2009; Levitt et al. 2000). It is well-known that neonates born small or preterm face greater risk of perinatal morbidity and mortality (Bernstein et al. 2000). Additionally, growing evidence suggests that LBW status also shapes health outcome across development into adulthood, affecting risk of a wide range of diseases such as diabetes (Harder et al. 2007; Whincup et al. 2008), hypertension (Mu et al. 2012), and neuroendocrinological (Jones et al. 2006; Wu□st et al. 2005) and inflammatory dysregulation (McDade et al. 2009). In turn, some of the long-term sequelae of being born small can increase the risk of giving birth to lighter babies in the next generation (Kuzawa & Sweet 2009; Thayer et al. 2012). These major public burdens caused by LBW have long impacted low- and middle-income countries (LMICs) at disproportionate levels (UNICEF-WHO 2019), which foreshadows a concerning public health burden caused by the increased risk of diseases associated with both LBW status.

A growing body of research has reported that gestating mothers who experience psychosocial stress are at heightened risk of giving birth to neonates with lower birthweights (Ae-Ngibise et al. 2019; Patil et al. 2020; Therrien et al. 2020; Zhu et al. 2010). Experiences of maternal prenatal stress are understood to contribute to LBW as a result of reduced gestational duration and restricted fetal growth, and often times, a complex combination of both gestational responses. Previous studies have shown that pregnant mothers with greater psychosocial stress tend to give birth to children with shorter gestational ages (Bussières et al. 2015; Coussons-Read 2012), who are preterm (Rosa et al. 2019; Wadhwa et al. 2011; Zhu et al. 2010), who are small-for-gestational age (SGA) (Class et al. 2011; Khashan et al. 2014), and in some cases, infants who are growth restricted (Berkowitz et al. 2003; Glynn et al. 2001; Resnik 2002). Additionally, previous studies have documented a wide variety of stressors that also predict LBW, which include pregnancy-related stress (Hobel et al. 2008), poverty (Lee & Lim 2010), racial discrimination (Collins et al. 2004), and maternal depression (Grote et al. 2010) and anxiety (Ding et al. 2014). Conversely, some researchers have found no evidence for impacts of stress on birthweight (Engel et al. 2005; Ramchandani et al. 2010).

While the mechanisms underlying the impacts of maternal cortisol on fetal development are only partially understood (Gitau et al. 2001), psychosocial stress exposure during earlier windows of gestation may lead to decreases in birthweight due to neuroendocrine-mediated fetal growth restriction and earlier parturition (D’Anna-Hernandez et al. 2012; Glynn et al. 2001). Three potential mechanisms have been suggested: maternal cortisol exposure through the placenta, downregulation of cortisol-shielding enzymatic mechanisms, and reduced stress reactivity across pregnancy. First, stress-initiated elevations in circulating glucocorticoids (GCs) may lead to greater levels of cortisol to move through the placenta and reach the fetus (O’Donnell et al. 2009). Greater maternal cortisol levels during early periods of pregnancy have been associated with being SGA (Goedhart et al. 2010) and LBW (Bolten et al. 2011; D’Anna-Hernandez et al. 2012; Goedhart et al. 2010). Contrary to the negative feedback loop between adrenal cortisol exposure and hypothalamic corticotropin-releasing hormone (CRH) production, higher cortisol exposure *increases* CRH production in the placenta, which subsequently can upregulate the production of pituitary adrenocorticotropic hormone (ACTH) and in turn, cortisol in both the mother and the fetus (Majzoub & Karalis 1999). Growing evidence also suggests that greater prenatal stress exposure may result in maternal immune dysregulation during pregnancy, which has been associated with adverse gestational development and birth outcomes, including restricted fetal growth, preterm birth, and LBW (Beijers et al. 2014; Nazzari et al. 2019).

Second, early increases in maternal cortisol during gestation may also downregulate fetal buffering mechanisms against maternal cortisol. Placental levels of 11β-hydroxysteroid dehydrogenase (11β-HSD-2) act as a natural shield against maternal cortisol *in utero* by converting cortisol into biologically inactive cortisone. Maternal cortisol, however, is still able to penetrate the fetus in small amounts particularly when circulating maternal cortisol reaches high concentrations or when 11β-HSD-2 is downregulated (O’Donnell et al. 2012). For instance, recent evidence reports that high levels of maternal GCs can consequently downregulate 11β-HSD-2 production, thus allowing greater passage of GCs, including cortisol, into the womb (O’Donnell et al. 2012). Placental 11β-HSD-2 levels are naturally and relatively low during early gestation (Hobel et al. 2008), and during this time cortisol may more easily pass through the placenta during earlier stages of pregnancy to slow fetal growth rate and shorten GA.

Finally, the impacts of early prenatal stressors on fetal development and birthweight may be heightened as behavioral and biological stress sensitivity decreases among mothers over the course of pregnancy. The sensitivity of both major stress-regulatory systems, the HPA and the sympathetic-adrenal-medullary (SAM) axes, declines as pregnancy advances. For example, mothers in advanced stages of pregnancy show decreased psychological sensitivity (Glynn et al. 2001), HPA axis reactivity (Kammerer et al. 2002; Obel et al. 2005), and decreased blood pressure responses to stress (Nisell et al. 1985) relative to those in earlier stages.

While the mechanisms by which maternal GCs contribute to intrauterine growth restriction and preterm birth are unclear, high levels of stress (Glynn et al. 2001; Lederman et al. 2004) and cortisol during the first trimester have consistently been shown to predict LBW (Bolten et al. 2011; D’Anna-Hernandez et al. 2012; Goedhart et al. 2010). This stands in contrast to the potential impacts of second or third trimester stress on LBW risk, which may play less of a role compared to first trimester stress due to the relative dampening of stress reactivity among late-stage pregnant women (Glynn et al. 2001; Kammerer et al. 2002) as well as the higher levels of 11β-HSD-2 buffering as pregnancy progresses. Thus, during times of low 11β-HSD-2 buffering, normal psychological and physiological stress reactivity, and high levels of stress-initiated maternal cortisol and placental CRH secretions *in utero*, cortisol may more easily pass through the placenta during the first trimester to affect the developing fetus to possibly slow fetal growth rate and shorten gestational length. We hypothesize that greater exposure to maternal stress during earlier periods of gestation will correspond with lower birthweights.

Notably, studies of prenatal stress and birthweight predominantly draw from samples in North America and Europe (Bussières et al. 2015), which leads to a research bias that overemphasizes psychosocial experiences and social-environmental conditions affecting mothers among high-income, Western populations. Consequently, it also overlooks LMICs that face greater burdens of LBW, such as South Africa where rates of LBW are relatively high (14.2%) and surprisingly close to the rates observed in more poverty-stricken countries in the sub-Saharan region (UNICEF-WHO 2019). In South Africa, LBW is the second leading cause of death in children under 5 years of age (Bradshaw et al. 2003). These trends have been tied to a long history of institutionalized segregation, poverty, and social marginalization characteristic of the apartheid regime, which actively discriminated against people of color, including but not limited to individuals categorized as Black, and continues to affect these communities. To address these gaps, here we examine the timing-specific effects of maternal stress during pregnancy on birthweight in a sample of women living in Soweto, South Africa.

## METHODS

### Study Setting

This study was nested within a large pregnancy cohort study (Soweto First 1000-Day Cohort; S1000), based in the [redacted] in Soweto, South Africa between 2013 and 2016. Under the 1960 Group Areas Act, the apartheid government segregated communities of color to live in demarcated areas, and Soweto was reserved for people classified as Black. Today, Soweto is a large peri-urban area of Johannesburg comprised of ethnically-, linguistically-, socioeconomically-diverse African communities (Alexander et al. 2013).

Overall, S1000 aimed to understand the complex associations between multiple maternal factors and fetal and infant outcomes in an urban African context, and to identify the levers that could optimize maternal and child health within the first the 1000 days; from conception to two years of age. Inclusion criteria were as follows: resident of the greater Soweto area, <20 weeks pregnant and no known diagnosis of epilepsy or diabetes at the time of recruitment, 18 years or older and pregnant with a singleton, naturally conceived pregnancy. A total of 657 women were included in the analytical sample, and 173 women were excluded from the final sample because of missing data for key variables. All women provided written informed consent prior to their inclusion in the pregnancy component of the study. Ethics approval was obtained from the [redacted] Research Ethics Committee (M120524].

### Demographic, Health and Socio-Economic Variables

Maternal demographic variables were collected by trained research staff using interviewer-administered questionnaires at the first pregnancy visit (<14 weeks GA) (Table 1). Parity was defined as the number of previous births at a gestational age of 24 weeks or more - regardless of whether the infant was born alive or was stillborn (0 = no previous births, 1 = one or more previous births). Smoking and/or chewing tobacco was reported at baseline. HIV-status was self-reported at each visit and confirmed using the results recorded in the participant’s antenatal clinic card. All HIV-positive participants in this study were receiving antiretrovirals. Household socio-economic status was assessed using an asset index which scored each participant according to the number of assets that they possessed out of a possible 11 (electricity, radio, television, refrigerator, mobile phone, personal computer, bicycle, motorcycle/scooter, car, agricultural land, farm animals). This was based on standard measures used in the Demographic and Health Surveys household questionnaire and has been extensively utilized in this setting (Kagura et al. 2016). Maternal education was defined according to the highest level of completion.

**Table 1.** Demographic characteristics, social experience, and birth outcomes

| Variables | n = 657 | % | Range |
| --- | --- | --- | --- |
| *Demographics* |  |  |  |
| Infant gender (% female) | 326 | 49.6 |  |
| Age (at enrollment) | 30 (5.8) |  | 18 - 44 |
| 18-24 | 142 | 21.6 |  |
| 25-29 | 183 | 27.9 |  |
| 30-34 | 169 | 25.7 |  |
| 35-44 | 163 | 24.8 |  |
| Marital status |  |  |  |
| Single | 403 | 61.3 |  |
| Married/Cohabiting | 254 | 38.7 |  |
| Educational attainment (% attended) |  |  |  |
| No school or primary school | 17 | 2.6 |  |
| Secondary school | 478 | 72.8 |  |
| Professional/teaching/university | 162 | 24.7 |  |
| Maternal body-mass index (BMI) | 28.2 |  | 15.8 – 60.6 |
| Primiparity (count) |  |  | 1-9 |
| 0 | 589 | 89.7 |  |
| ≥1 | 68 | 10.3 |  |
| Smoking |  |  |  |
| No | 602 | 91.6 |  |
| Yes | 55 | 8.4 |  |
| *Social experience* |  |  |  |
| Prenatal stress, 1<sup>st</sup> trimester (count) | 4 (2.3) |  | 0-12 |
| Prenatal stress, 3<sup>rd</sup> trimester (count)* | 3 (2.0) |  | 0-9 |
| Social support, 1<sup>st</sup> trimester (count) | 6 (1.2) |  | 2-8 |
| Social support, 3<sup>rd</sup> trimester (count)* | 6 (1.1) |  | 2-7 |
| Maternal depression, 1<sup>st</sup> trimester (EPDS) | 9.4 (5.9) |  | 0-27 |
| Maternal anxiety, 1<sup>st</sup> trimester (STAI) | 9.4 (2.5) |  | 6-21 |
| Maternal depression, 1<sup>st</sup> trimester (EPDS) | 8.6 (5.8) |  | 0-26 |
| Maternal anxiety, 3<sup>rd</sup> trimester (STAI) | 9.6 (2.6) |  | 6-24 |
| *Infant birth outcomes* |  |  |  |
| Birth weight (g) | 2971 (578) |  | 730 – 4500 |
| Low birth weight (<2500g) | 109 | 16.6% |  |
| Gestational age (week) | 38.1 (2.4) |  | 25 - 42 |
| Preterm birth | 96 | 14.6% |  |

\*Due to attrition, the sample size for data collected during the 3<sup>rd</sup> trimester visit was 510.

### Anthropometry

A wall-mounted Stadiometer (Holtain, UK) was used to measure maternal height to the nearest 1 mm at baseline. Maternal weight was measured to the nearest 0.1 kg at each visit during pregnancy using a digital scale. Weight at recruitment (<14 weeks) was used as a proxy for pre-pregnancy weight and, together with height, was used to calculate maternal body mass index (BMI) (weight (kg)/height (m^2^)).

### Maternal prenatal stress and social support

A 14-item scale was used to assess stress (including relationship, family, economic, and societal stressors) during recruitment (<14 weeks). Each question records a yes/no response to whether the stressor of question was experienced in the past six months. This scale was developed in previous cohort work at [redacted] and is considered a reliable measure of stress (Ramchandani et al. 2010). While each scale assesses retrospective exposures of stress within the past six months, we treat each time of collection as the approximate time of antenatal stress exposure. Social support was measured using a series of nine questions to identify the absence or presence of instrumental and emotional support, including: people available to help, a confidante, being able to speak to her partner, belonging to a community organisation/ church and having a friend with a baby.

### Birth outcomes

Gestational age at delivery (weeks) was calculated as: [duration of pregnancy follow-up (date of delivery – date of baseline ultrasound dating scan) + GA at baseline (days)]. Birthweight was measured by trained research nurses within 24 hours of delivery for 82% of neonates. Where assessment within this window was not possible, measurements were taken within 48 hours.

### Statistical analyses

All variables were examined for normal distribution and outliers. Bivariate analyses were conducted between exposure variables, birthweight, and covariates. Covariates were included based on *a priori* knowledge of social, biological, and obstetric risk factors that may potentially confound the relationship between prenatal stress and birthweight. The following variables were considered for inclusion as confounding factors: marital status (single, married), household assets (number of assets owned among the following list of items: electricity, radio, television, refrigerator, mobile phone, personal computer, bicycle, motorcycle/scooter, car, agricultural land, and farm animals), maternal education (last year of education completed), household density (number of inhabitants divided by number of rooms available for sleeping), social support (composite score), alcohol consumption (yes/no), maternal depression (Edinburgh Postnatal Depression Scale) (Cox et al. 1996), maternal anxiety (State-Trait Anxiety Inventory) (Marteau & Bekker 1992), gestational diabetes (self-reported diagnostic status), and HIV status (self-reported) (*p* > 0.10).

Bivariate analyses were conducted to identify potential confounding variables for inclusion in the final analytic models (Table 2). With the exception of known confounding factors for birthweight, only those that were statistically significant at the 0.1 level were included the final models. Multiple ordinary least squares (OLS) regressions were conducted to examine the impact of prenatal stress on birthweight. Maximum likelihood ratio tests assessed whether the models with the interaction terms were an improvement over the main effects models at *p* < 0.15 for declaring the interaction term significant.

**Table 2.** Multiple regression models of first trimester prenatal stress scores predicting birthweight (grams)

|  | Model 1 | Model 2 | Model 3 | Model 4 | Model 5 | Model 6 | Model 7 |
| --- | --- | --- | --- | --- | --- | --- | --- |
| Prenatal stress | -7.6 ± 10.0 | -7.2 ± 10.0 | -7.0 ± 9.9 | -6.3 ± 9.8 | -12.4 ± 7.1^ | -12.7 ± 7.0^ | -10.3 ± 7.2 |
| Gender (female) |  | -76.4 ± 45.1^ | -72.7 ± 45.0 | -76.1 ± 44.4^ | -114.1 ± 32.0*** | -109.8 ± 31.8** | -109.3 ± 31.8** |
| Maternal age |  |  | -7.1 ± 3.9^ | -9.0 ± 3.8* | -3.0 ± 2.8 | -5.1 ± 2.8^ | -5.3 ± 2.8^ |
| Maternal BMI |  |  |  | 16.1 ± 3.6*** | 15.0 ± 2.6*** | 14.0 ± 2.6*** | 13.9 ± 2.6*** |
| Gestational age |  |  |  |  | 23.7 ± 1.0*** | 24.0 ± 1.0*** | 24.0 ± 1.0*** |
| Primiparity |  |  |  |  |  | -165.7 ± 54.7** | -158.1 ± 54.9** |
| Smoking |  |  |  |  |  |  | -94.5 ± 59.3 |
| Intercept | 2997.5 ± 42.9*** | 3034.0 ± 47.9*** | 3244.7 ± 124.1*** | 2849.1 ± 151.0*** | -3658.9 ± 285.7*** | -3635.6 ± 284.0*** | -3616.1 ± 284.0*** |
| Model R^2 | 0.0009 | 0.0053 | 0.0104 | 0.0399 | 0.5034 | 0.5103 | 0.5122 |

^*p* < 0.10 \**p* < 0.05 \*\**p* < 0.01 \*\*\**p* < 0.001

**Table 2a.** Zero-order correlations across study variables

|  | 1 | 2 | 3 | 4 | 5 | 6 | 7 | 8 |
| --- | --- | --- | --- | --- | --- | --- | --- | --- |
| 1. Birthweight | 1 |  |  |  |  |  |  |  |
| 2. Stress | 0.0299 | 1 |  |  |  |  |  |  |
| 3. Gender | -0.0668 | 0.0222 | 1 |  |  |  |  |  |
| 4. Maternal age | -0.0749 | 0.0122 | 0.0455 | 1 |  |  |  |  |
| 5. Maternal BMI | 0.1615 | -0.0142 | 0.0221 | 0.1121 | 1 |  |  |  |
| 6. Gestational age | 0.6822 | 0.0346 | 0.0451 | -0.0846 | 0.0073 | 1 |  |  |
| 7. Primiparity | -0.0125 | -0.0081 | 0.0331 | -0.2654 | -0.1470 | 0.1242 | 1 |  |
| 8. Smoking | -0.0719 | 0.2071 | 0.0138 | -0.0644 | -0.0445 | -0.0083 | 0.0983 | 1 |

\* $p < 0.05$

## RESULTS

This study included 657 mother and infants (Table 1). Participants included in the analytical sample were similar to those excluded (n = 173) with respect to first trimester prenatal stress, birthweight, fetal gender, maternal BMI, and GA (*p* > 0.05). Late pregnancy prenatal stress, primiparity, and maternal age were significantly different from those excluded from the sample (*p* < 0.05). Mothers in the analytical sample exhibited slightly lower average levels of stress, reported marginally higher rates of primiparity, and were older by about one year. Fetal gender, maternal age, maternal BMI, primiparity, and GA were significantly associated with birthweight.

Table 3 presents the results of the OLS regression analyses of maternal and fetal factors that predict infant birthweight. The unadjusted model (Model l) predicting birthweight on first trimester prenatal stress, while insignificant, displays a negative relationship as expected. This relationship between prenatal stress and birthweight remains insignificant after adjusting for fetal gender, maternal age, and maternal BMI (Model 2 & 3). The association between prenatal stress approaches significance (β = −12.4, *p* = 0.079, 95% CI [-26.3, 1.44]) after controlling for GA, which strongly predicts birthweight and accounts for a large portion of the variation (46%). Finally, prenatal stress remains inversely related to and a modest predictor of birthweight after adjusting for primiparity. A one-point increase in prenatal stress scores corresponded with a - 12.7 g decrease in birthweight (β = −12.7, *p* = 0.071, 95% CI [-26.4, 1.10]). Gender, maternal BMI, GA, and primiparity significantly relate to birthweight and maternal age serves as a modest predictor of birthweight. After adding maternal smoking to the model (Model 7), the effect of prenatal stress strengthens but becomes insignificant. The interaction between prenatal stress and gender was not significant (*p* = 0.752, 95% CI [-32.1, 23.2]) suggesting that there were no gender differences in the effects of prenatal stress on birthweight in our sample. An identical logistic model predicting LBW was also analyzed and found that the effect of prenatal stress on LBW was non-significant yet trended in the expected direction (*p* = 0.673, OR = 1.03, 95% CI [0.90, 1.17]).

The effect of first trimester maternal stress on GA was also examined to determine whether shorter gestational periods contributed to lower birthweights. Both unadjusted and fully adjusted models show that the effect of first trimester prenatal stress is not associated with GA (results not shown). Specifically, these analyses also reported a positive but non-significant relationship between maternal stress and GA: women with higher levels of maternal stress during their first trimester seemed to have longer gestations.

## DISCUSSION

We report an inverse relationship between maternal stress severity and infant birthweight: first trimester maternal stress is associated with lower birthweights in our sample of women in Soweto, South Africa. There was also no evidence that the relationship between stress and birthweight was modified by maternal depression, anxiety, and perceived social support. These results point to the importance of the first trimester as an important developmental period for infant birth outcomes.

Previous studies have also reported the inverse relationship between higher first trimester prenatal stress and lower birthweights (Coussons-Read et al. 2012; Dancause et al. 2011; Ryu 2019; Vrijkotte et al. 2009; Zhu et al. 2010). For example, in a large sample of expectant mothers in Hefei, China, self-reported stressful life events that occurred during the first trimester predicted lower birthweights, which were unrelated to stress during the second or third trimester (Zhu et al. 2010). Additionally, in a study of the impacts of a severe ice storm, GA and birthweights were lower among women exposed to the ice storm during early and mid-pregnancy (Dancause et al. 2011). Conversely, a number of studies have also reported no evidence for the effects of first trimester stress on birthweight (Rondó et al. 2003). A wide range of severity and type of maternal stress exposure have been examined in past literature, (e.g. pregnancy-related distress, natural disasters, domestic violence, poverty, trauma, etc.) and the downstream biological pathways linking certain forms of maternal experience and LBW may vary, such as the case of maternal depression (Field et al. 2006). Nevertheless, the larger relationship between maternal prenatal stress that occurred during the first trimester and infant birthweight is consistently documented across study populations.

Our final model (Model 7) also uncovered the deleterious effect of maternal smoking during pregnancy on lowering infant birthweights. While the effect of maternal stress approached significance after controlling for key covariates, the prenatal stress variable became statistically insignificant after including the smoking variable into the model, which itself was not a significant predictor of birthweight. Maternal smoking during pregnancy is a well-known risk factor for LBW (Magee et al. 2004; Steyn et al. 2006) and is understood to lower birthweight through various pathways, including fetal hypoxia (Abel 1980), uterine vasoconstriction (Quigley et al. 1979), and oxidative stress-mediated alterations in placental function (Stone et al. 2014), all of which can contribute to fetal growth restriction. Further analyses indicate that prenatal stress and smoking are collinear with one another (results not shown), suggesting a possible bi-directional relationship between the two variables: smokers may exhibit higher stress levels than non-smokers, or highly stressed women may be more likely to smoke as a coping mechanism (Lobel et al. 2008). Because of this collinearity, our measure of prenatal stress alone may adequately capture the negative impacts of both stress and smoking on infant birthweight.

While GA was a significant and expected predictor of birthweight and accounted for 46% of the variation in our models, contrary to the larger literature (Glynn et al. 2008; Zhu et al. 2010), prenatal stress does not appear to lower birthweight as a result of shortened GA in this sample. Our data reported a positive but statistically insignificant relationship between the severity of stress exposure during early pregnancy and GA. It is only after controlling for gestational duration that the negative association between early prenatal stress and birthweight approaches significance (Model 6). This pattern suggests that fetal growth is likely slowed in relation to first trimester stress, but that this stress-linked delay in fetal growth is masked by non-significantly longer gestations among women in our sample. The lack of an association between stress and GA may be explained by prior global epidemiological patterns of birth outcomes by location, which show that in high-income countries, the majority of LBW babies are the result of shortened gestation rather than growth restriction (Mattison et al. 2001). In contrast, in LMICs, LBW persists irrespective of GA as well as an increased prevalence of SGA due to environmental and nutritional conditions such as greater infectious disease burden and undernutrition, which compromise maternal metabolism and the overall development of the fetus (Pike 2005).

While the prenatal stress scale was operationalized as a marker of maternal stress at the time of collection, the timing effects may reasonably reflect exposures experienced up to six months prior to data collection as the scale assessed retrospective maternal exposures within the past six months. Because the stress assessment was administered before 14 weeks of pregnancy, the measure may represent experiences up to three to six months before pregnancy. Pre-pregnancy maternal stress has also predicted LBW in past studies (Khashan et al. 2008). In the present study, there is also no evidence for the protective effects of social support as the main effects of social support and the interaction between stress and social support was not significant.

In summary, women who experienced psychosocial stress early in gestation gave birth to modestly lower birthweight babies in our sample of South African mothers and infants. Women who experienced greater levels of stress appeared to have non-significantly longer gestations, and the negative impact of maternal stress on infant birthweight only appeared after adjusting for this pattern. These results suggest that delays in fetal growth associated with first trimester maternal stress may contribute to lowering birthweights in our sample. Our findings emphasize the importance of better understanding the sources of stress experienced among women of reproductive age in Soweto in order create social interventions aimed at reducing maternal stress, especially early in gestation. Furthermore, this study informs larger public health campaigns, such as earlier antenatal visits, earlier ultrasound screenings, and better monitoring of maternal psychosocial states during pregnancy, designed to ameliorate and prevent the high morbidity of poor birth outcomes across South Africa and other LMICs.

## Data Availability

Data are available upon request from the Developmental Pathways for Health Research Unit research team.

## Acknowledgements

We are indebted to the research participants who took part in this study and our research assistants who worked tirelessly to make this project possible. This work would not be possible without their effort.

## Bibliography

Abel, E. L. (1980). Smoking during pregnancy: a review of effects on growth and development of offspring. Human biology, 593–625.

Ae-Ngibise, K., Jack, D., Wylie, B., Oppong, F., Kaali, S., Agyei, O., Kinney, P., Wright, R., Asante, K., Lee, A. and GRAPHS study team. (2019). Impact of prenatal maternal stress on birth anthropometrics and pregnancy outcomes in rural Ghana. Environmental Epidemiology, 3, 4.

Alexander, P., Ceruti, C., Motseke, K., Phadi, M., & Wale, K. (2013). Class in Soweto. University of KwaZulu-Natal Press.

Barker, D. J. (1998). In utero programming of chronic disease. Clinical science, 95(2), 115–128.

Beijers, R., Buitelaar, J. K., & de Weerth, C. (2014). Mechanisms underlying the effects of prenatal psychosocial stress on child outcomes: beyond the HPA axis. European child & adolescent psychiatry, 23(10), 943–956.

Berghänel, A., Heistermann, M., Schülke, O., & Ostner, J. (2017). Prenatal stress accelerates offspring growth to compensate for reduced maternal investment across mammals. Proceedings of the National Academy of Sciences, 114(50), E10658–E10666.

Berkowitz, G. S., Wolff, M. S., Janevic, T. M., Holzman, I. R., Yehuda, R., & Landrigan, P.J. (2003). The World Trade Center disaster and intrauterine growth restriction. Jama, 290(5), 595–596.

Bradshaw, D., Bourne, D., & Nannan, N. (2003). What are the leading causes of death among South African children. MRC policy brief, 3, 1–4.

Bolten, M. I., Wurmser, H., Buske-Kirschbaum, A., Papoušek, M., Pirke, K. M., & Hellhammer, D. (2011). Cortisol levels in pregnancy as a psychobiological predictor for birthweight. Archives of women’s mental health, 14(1), 33–41.

Bussieres, E. L., Tarabulsy, G. M., Pearson, J., Tessier, R., Forest, J. C., & Giguere, Y. (2015). Maternal prenatal stress and infant birthweight and gestational age: A meta-analysis of prospective studies. Developmental Review, 36, 179–199.

Class, Q. A., Lichtenstein, P., Långström, N., & D’onofrio, B. M. (2011). Timing of prenatal maternal exposure to severe life events and adverse pregnancy outcomes: a population study of 2.6 million pregnancies. Psychosomatic medicine, 73(3), 234.

Collins J. W., David, R. J., Handler, A., Wall, S., & Andes, S. (2004). Very low birthweight in African American infants: the role of maternal exposure to interpersonal racial discrimination. American journal of public health, 94(12), 2132–2138.

Coussons-Read, M. E., Lobel, M., Carey, J. C., Kreither, M. O., D’Anna, K., Argys, L., Ross, R.G., Brandt, C. & Cole, S. (2012). The occurrence of preterm delivery is linked to pregnancy-specific distress and elevated inflammatory markers across gestation. Brain, behavior, and immunity, 26(4), 650–659.

Cox, J. L., Chapman, G., Murray, D., & Jones, P. (1996). Validation of the Edinburgh Postnatal Depression Scale (EPDS) in non-postnatal women. Journal of affective disorders, 39(3), 185–189.

D’Anna-Hernandez, K. L., Hoffman, M. C., Zerbe, G. O., Coussons-Read, M., Ross, R. G., & Laudenslager, M. L. (2012). Acculturation, maternal cortisol and birth outcomes in women of Mexican descent. Psychosomatic Medicine, 74(3), 296.

Diego, M.A., Field, T., Hernandez-Reif, M., Schanberg, S., Kuhn, C., Gonzalez-Quintero, V.H., 2009. Prenatal depression restricts fetal growth. Early Human Development. 85, 65–70. Doi:10.1016/j.earlhumdev.2008.07.002.

Ding, X.X., Wu, Y.L., Xu, S.J., Zhu, R.P., Jia, X.M., Zhang, S.F., Huang, K., Zhu, P., Hao, J.H., & Tao, F.B. (2014). Maternal anxiety during pregnancy and adverse birth outcomes: a systematic review and meta-analysis of prospective cohort studies. Journal of affective disorders, 159, 103–110.

Engel, S. M., Berkowitz, G. S., Wolff, M. S., & Yehuda, R. (2005). Psychological trauma associated with the World Trade Center attacks and its effect on pregnancy outcome. Paediatric and perinatal epidemiology, 19(5), 334–341.

Field, T., & Diego, M. (2008). Cortisol: the culprit prenatal stress variable. International Journal of Neuroscience, 118(8), 1181–1205.

Field, T., Diego, M., & Hernandez-Reif, M. (2006). Prenatal depression effects on the fetus and newborn: a review. Infant Behavior and Development, 29(3), 445–455.

Gitau, R., Fisk, N. M., & Glover, V. (2001). Maternal stress in pregnancy and its effect on the human foetus: an overview of research findings. Stress, 4(3), 195–203.

Glover, V., Bergman, K., Sarkar, P., & O’Connor, T. G. (2009). Association between maternal and amniotic fluid cortisol is moderated by maternal anxiety. Psychoneuroendocrinology, 34(3), 430–435.

Glover, V., & O’Connor, T. G. (2002). Effects of antenatal stress and anxiety: implications for development and psychiatry. The British Journal of Psychiatry, 180(5), 389–391.

Glynn, L. M., Wadhwa, P. D., Dunkel-Schetter, C., Chicz-DeMet, A., & Sandman, C. A. (2001). When stress happens matters: effects of earthquake timing on stress responsivity in pregnancy. American journal of obstetrics and gynecology, 184(4), 637–642.

Glynn, L. M., Schetter, C. D., Hobel, C. J., & Sandman, C. A. (2008). Pattern of perceived stress and anxiety in pregnancy predicts preterm birth. Health Psychology, 27(1), 43.

Goedhart, G., Vrijkotte, T. G., Roseboom, T. J., van der Wal, M. F., Cuijpers, P., & Bonsel, G. J. (2010). Maternal cortisol and offspring birthweight: results from a large prospective cohort study. Psychoneuroendocrinology, 35(5), 644–652.

Grote, N. K., Bridge, J. A., Gavin, A. R., Melville, J. L., Iyengar, S., & Katon, W. J. (2010). A meta-analysis of depression during pregnancy and the risk of preterm birth, low birthweight, and intrauterine growth restriction. Archives of general psychiatry, 67(10), 1012–1024.

Harder, T., Rodekamp, E., Schellong, K., Dudenhausen, J. W., & Plagemann, A. (2007). Birthweight and subsequent risk of type 2 diabetes: a meta-analysis. American journal of epidemiology, 165(8), 849–857.

Hobel, C. J., Goldstein, A. M. Y., & Barrett, E. S. (2008). Psychosocial stress and pregnancy outcome. Clinical obstetrics and gynecology, 51(2), 333–348.

Jones, A., Godfrey, K. M., Wood, P., Osmond, C., Goulden, P., & Phillips, D. I. (2006). Fetal growth and the adrenocortical response to psychological stress. The Journal of Clinical Endocrinology & Metabolism, 91(5), 1868–1871.

Khashan, A. S., Everard, C., McCowan, L. M. E., Dekker, G., Moss-Morris, R., Baker, P. N., et al. (2014). Second-trimester maternal distress increases the risk of small for gestational age. Psychological Medicine, 44, 1–12.

Khashan, A. S., McNamee, R., Abel, K. M., Pedersen, M. G., Webb, R. T., Kenny, L. C., et al. (2008). Reduced infant birthweight consequent upon maternal exposure to severe life events. Psychosomatic Medicine, 70, 688–694.

Kramer MS. 1987. Determinants of low birthweight: methodological assessment and meta-analysis. Bull WHO 65:663–737.

Kuzawa, C. W., & Sweet, E. (2009). Epigenetics and the embodiment of race: developmental origins of US racial disparities in cardiovascular health. American Journal of Human Biology: The Official Journal of the Human Biology Association, 21(1), 2–15.

Lederman, S. A., Rauh, V., Weiss, L., Stein, J. L., Hoepner, L. A., Becker, M., & Perera, F. P. (2004). The effects of the World Trade Center event on birth outcomes among term deliveries at three lower Manhattan hospitals. Environmental health perspectives, 112(17), 1772–1778.

Lee, B. J., & Lim, S. H. (2010). Risk of low birthweight associated with family poverty in Korea. Children and Youth Services Review, 32(12), 1670–1674.

Levitt, N. S., Lambert, E. V., Woods, D., Hales, C. N., Andrew, R., & Seckl, J. R. (2000). Impaired glucose tolerance and elevated blood pressure in low birthweight, nonobese, young South African adults: early programming of cortisol axis. The Journal of Clinical Endocrinology & Metabolism, 85(12), 4611–4618.

Lobel, M., Cannella, D. L., Graham, J. E., DeVincent, C., Schneider, J., & Meyer, B. A. (2008). Pregnancy-specific stress, prenatal health behaviors, and birth outcomes. Health Psychology, 27, 604–615.

Magee, B. D., Hattis, D., & Kivel, N. M. (2004). Role of smoking in low birthweight. The Journal of reproductive medicine, 49(1), 23–27.

Marteau, T. M., & Bekker, H. (1992). The development of a sixLJitem shortLJform of the state scale of the Spielberger State—Trait Anxiety Inventory (STAI). British journal of clinical Psychology, 31(3), 301–306.

Mattison D, Damus K, Fiore E, Petrini J, Alter C. 2001. Preterm delivery: a public health perspective. Pediat Perinat Epidemiol 15:7–16.

Majzoub, J. A., & Karalis, K. P. (1999). Placental corticotropin-releasing hormone: function and regulation. American journal of obstetrics and gynecology, 180(1), S242–S246.

McDade, T. W., Rutherford, J., Adair, L., & Kuzawa, C. W. (2009). Early origins of inflammation: microbial exposures in infancy predict lower levels of C-reactive protein in adulthood. Proceedings of the Royal Society B: Biological Sciences, 277(1684), 1129–1137.

Mu, M., Wang, S. F., Sheng, J., Zhao, Y., Li, H. Z., Hu, C. L., & Tao, F. B. (2012). Birthweight and subsequent blood pressure: a meta-analysis. Archives of cardiovascular diseases, 105(2), 99–113.

Murphy, V. E., & Clifton, V. L. (2003). Alterations in human placental 11β-hydroxysteroid dehydrogenase type 1 and 2 with gestational age and labour. Placenta, 24(7), 739–744.

Obel, C., Hedegaard, M., Henriksen, T. B., Secher, N. J., Olsen, J., & Levine, S. (2005). Stress and salivary cortisol during pregnancy. Psychoneuroendocrinology, 30(7), 647–656.

O’Donnell, K., O’Connor, T. G., & Glover, V. (2009). Prenatal stress and neurodevelopment of the child: focus on the HPA axis and role of the placenta. Developmental neuroscience, 31(4), 285–292.

O’Donnell, K. J., Jensen, A. B., Freeman, L., Khalife, N., O’Connor, T. G., & Glover, V. (2012). Maternal prenatal anxiety and downregulation of placental 11β-HSD2. Psychoneuroendocrinology, 37(6), 818–826.

Pagel, M. D., Smilkstein, G., Regen, H., & Montano, D. (1990). Psychosocial influences on new born outcomes: a controlled prospective study. Social science & medicine, 30(5), 597–604.

Patil, D., Enquobahrie, D. A., Peckham, T., Seixas, N., & Hajat, A. (2020). Retrospective cohort study of the association between maternal employment precarity and infant low birth weight in women in the USA. BMJ open, 10(1).

Quigley, M. E., Sheehan, K. L., Wilkes, M. M., & Yen, S. S. C. (1979). Effects of maternal smoking on circulating catecholamine levels and fetal heart rates. American Journal of Obstetrics and Gynecology, 133(6), 685–690.

Ramchandani, P. G., Richter, L. M., Norris, S. A., & Stein, A. (2010). Maternal prenatal stress and later child behavioral problems in an urban South African setting. Journal of the American Academy of Child & Adolescent Psychiatry, 49(3), 239–247.

Resnik, R. (2002). Intrauterine growth restriction. Obstetrics & Gynecology, 99(3), 490–496.

Rondó, P. H., Ferreira, R. F., Nogueira, F., Ribeiro, M. C., Lobert, H., & Artes, R. (2003). Maternal psychological stress and distress as predictors of low birthweight, prematurity and intrauterine growth retardation. European journal of clinical nutrition, 57(2), 266.

Rosa, M. J., Nentin, F., Bosquet Enlow, M., Hacker, M. R., Pollas, N., Coull, B., & Wright, R. J. (2019). Sex-specific associations between prenatal negative life events and birth outcomes. Stress, 22(6), 647–653.

Ryu, H. (2019). Maternal prenatal stress and birth weight (Doctoral dissertation, KDI School).

Steyn, K., De Wet, T., Saloojee, Y., Nel, H., & Yach, D. (2006). The influence of maternal cigarette smoking, snuff use and passive smoking on pregnancy outcomes: the Birth To Ten Study. Paediatric and perinatal epidemiology, 20(2), 90–99.

Stone, W. L., Bailey, B., & Khraisha, N. (2014). The pathophysiology of smoking during pregnancy: a systems biology approach. Front Biosci (Elite Ed), 6(2), 318–328.

Thayer, Z. M., Feranil, A. B., & Kuzawa, C. W. (2012). Maternal cortisol disproportionately impacts fetal growth in male offspring: evidence from the Philippines. American Journal of Human Biology, 24(1), 1–4.

Therrien, A.S., Buffa, G., Roome, A., Standard, E., Taleo, G., Tarivonda, L., Olszowy, K.M. and Dancause, K.N. (2020, April). Prenatal stress predicts birthweight independent of maternal dietary patterns: results of a prospective longitudinal study in Vanuatu. In AMERICAN JOURNAL OF HUMAN BIOLOGY (Vol. 32). 111 RIVER ST, HOBOKEN 07030-5774, NJ USA: WILEY.

Vrijkotte, T. G., Van Der Wal, M. F., Van Eijsden, M., & Bonsel, G. J. (2009). First-trimester working conditions and birthweight: a prospective cohort study. American journal of public health, 99(8), 1409–1416.

Wadhwa, P. D., Entringer, S., Buss, C., & Lu, M. C. (2011). The contribution of maternal stress to preterm birth: issues and considerations. Clinics in perinatology, 38(3), 351-384.2

Whincup, P.H., Kaye, S.J., Owen, C.G., Huxley, R., Cook, D.G., Anazawa, S., Barrett-Connor, E., Bhargava, S.K., Birgisdottir, B.E., Carlsson, S., & De Rooij, S.R (2008). Birthweight and risk of type 2 diabetes: a systematic review. JAMA, 300(24), 2886–2897.

World Health Organization. (2019). UNICEF-WHO low birthweight estimates: levels and trends 2000-2015 (No. WHO/NMH/NHD/19.21). United Nations Children’s Fund (UNICEF).

Zhu, P., Tao, F., Hao, J., Sun, Y., & Jiang, X. (2010). Prenatal life events stress: implications for preterm birth and infant birthweight. American journal of obstetrics and gynecology, 203(1), 34–e1.

